# Systematic druggable genome-wide Mendelian randomization identifies therapeutic target genes for chronic periodontitis

**DOI:** 10.1101/2025.01.19.25320804

**Authors:** Zhongwei Cheng, Fang Wang, Meimei Ran, Shenhu Liang, Qinggao Song

**Affiliations:** Department of Oral and Maxillofacial Surgery Affiliated Stomatological Hospital of Zunyi Medical University, Zunyi, Guizhou, China; School and Hospital of Stomatology, Zunyi Medical University, Zunyi, Guizhou, China; Information Technology Network Management Center,Zunyi Medical And Pharmaceutical College, Zunyi, Guizhou, China

**Keywords:** Chronic periodontitis, Genome-wide Mendelian randomisation, Target genes

## Abstract

Currently,the treatment of chronic periodontitis (CP) still has challenges. This study aimed to identify novel drug targets for the treatment of chronic periodontitis in the druggable genome using the Mendelian randomization (MR) method.In this study,based on the list of 4479 drug gene targets,overlapping genes were selected in blood expression Quantitative Trait Loci(eQTL), which were then subjected to Two-Sample MR(TSMR) and validated Fusion Transcriptome-Wide Association Study(TWAS) with data from the Genome-Wide Association Study(GWAS) of CP to confirm drug genes genetically associated with CP and tested for multiplicity of effects using Summary-data-based Mendelian randomization(SMR) analysis and colocalisation. Finally, Phenome-Wide association study(PheWAS) was executed on the identified drug targets.We applied SMR,TSMR,Fusion TWAS,and a series of manners of co-localisation to assess the genetic association between drugable targets and CP. In conclusion,Metalloproteinase 25(MMP25)was considered the most promising drug target,in addition to which we maintained some confidence in TNFRSF18,CDC25B,STK10 and ACVR2B.Finally,the PheWAS-MR results showed that the possible side effects of applying MMP25 inhibitors were Otitis externa as well as some metabolic disorders,among others.In summary,we identified five potential CP drug targets using TSMR,Fusion TWAS,SMR,and co-localisation a series of methods,of which MMP25 passed all the tests.Such discovery offered an academic basis for future CP drug development and reduce drug development time and economic costs to some extent.

## Introduction

Periodontitis affects approximately 70-80% of adults and is the sixth most epidemic illness around the world [1], it causes severe inflammation in the tissues surrounding teeth, such as gums, alveolar bone and periodontal ligament, which, in addition to the tooth loss, bone loss, and formation of periodontal pockets [2,3], is also closely associated with a number of systemic illness, such as diabetes mellitus, cardiovascular disease, and chronic kidney disease [4-7]. The persistent progression of gingival recession in chronic periodontitis (CP) severely reduces the patients’ life quality and brings social health organisations a great challenge [1]. In fact, the pathogenesis of chronic periodontitis is complex and multifactorial, and although microbial infection of the periodontal biofilm is the main etiological factor, it is also largely related to a combination of host autoimmune characteristics, environmental factors, and genetic susceptibility[8-10], which makes it difficult to treat chronic periodontitis effectively.

Previous studies have shown that anti-cytokine therapy (inhibition of tumour necrosis factor, interleukin-1 or interleukin-17 action) has demonstrated some efficacy in experimental models of periodontitis [11] and several clinical studies have evaluated antirheumatic drugs (anabolic acid, etanercept and infliximab, among others) and NSAIDs as having a positive effect on periodontitis [12]. In addition, Amyndas Pharmaceuticals approved the development of new drugs (e.g. improved analogues of the C3 inhibitor conistatin family, i.e. Cp40), which set the stage for the progression of AMY-101 [13-15]. However, in host regulation therapies, there may be potential safety problems involving biological targeting specific components of the immune response, includes the infection risk increased, especially with the long-term use of NSAIDs, where the possibility of ill effects and unknown hazards is greater [9][16,17]. Therefore, there is a need to continue to explore potential therapeutic gene targets for CP therapy.

Mendelian randomization (MR) has been used in various fields of medicine as an effective method to speculate causality between two phenotypes by genetic variants as instrumental variables (IVs) [18]. In addition MR analysis can be used to find new drug targets by consolidating pooled data from illness genome-wide association studies (GWAS) and expression quantitative trait loci (eQTL) researchs [19]. In druggable genes’ genomic regions, eQTLs was found to represent characterising traits, as the level of gene expression could be viewed as a kind of lifelong exposure [20]. So we conducted a scientific genome-wide MR study to search possible targets for chronic periodontitis treatment. Firstly, we screened cis-eQTLs from blood eQTLs through information of drug genes list. Using GWAS data of chronic periodontitis, two-sample MR analysis was taked to identify genes highly related to chronic periodontitis. Then we performed SMR analysis, co-localisation analysis, and TWAS analysis to prove the robustness of the MR results. Phenome-wide association study (PheWAS) was then performed on the identified drug targets.

## Materials and methods

Our study design process is shown in Fig1.The following is a detailed description of the methods and materials used in this study.

**Figure 1.**
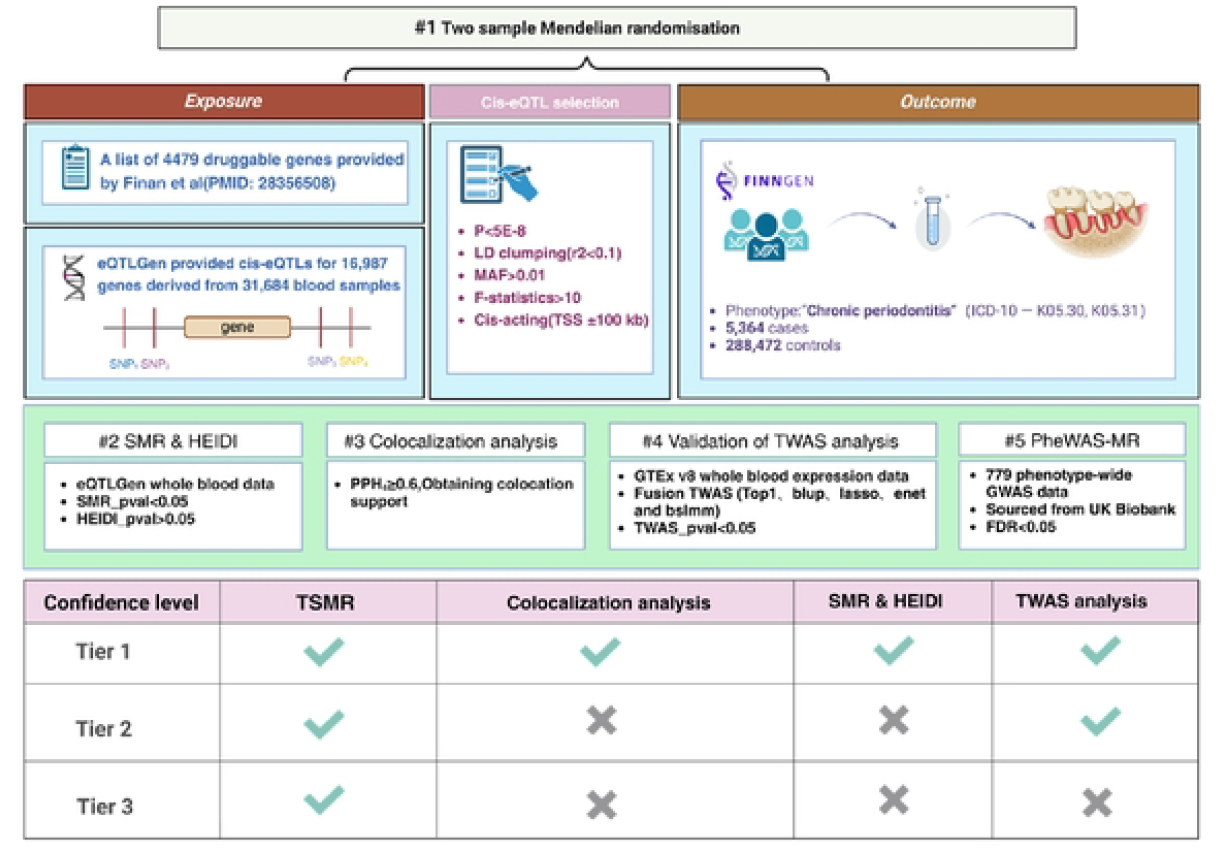
Flowchart of the design of this study. MAF: Minor allele frequency; eQTL: Expression quantitative trait loci; SMR:Summary-data-based Mendelian randomization;GWAS: Genome-wide association Studies; TWAS:Transcriptome-wide association analysis; PheWAS: Phenome-wide association study.

### Source of druggable gene targets

In order to be able to provide a comprehensive and credible list of druggable gene targets for subsequent studies, we selected a list of 4,479 druggable genes (20,300 protein-coding genes annotated in Ensembl v.73) consolidated by Finan C, et al [21], which has 31.9% (1,427/4479) of genes with approved or clinical-stage drug targets, 15.2% (689/4479) of the genes are similar to approved drug targets and 52.9% (2370/4479) encode proteins that are remotely similar to approved drug targets, which are widely used in many studies today [22]. Information of druggable genes on the list comes from the original article’s publicly available supplementary material.

### Chronic periodontitis GWAS data and eQTL dataset GWAS data sources

The raw eQTL public data downloaded from the official eQTLGen (https://eqtlgen.org/) website [23], which summarises cis-eQTL for 16,987 genes (318,614 blood samples from healthy individuals of European descent), and the current study only included samples from this raw dataset with false discovery rate (FDR) < 0.05 in this original dataset.

Chronic periodontitis GWAS data were obtained from the Finnish database version R11 (updated 24 June 2024), which includes 5,364 cases and 288,472 controls of European origin, in the Finnish database under the category ‘Diseases of the digestive system’. The phenotype ‘Chronic periodontitis’ was searched in the Finnish database under the category ‘Diseases of the digestive system’ and was downloaded with reference to ICD-10:K05.30 and K05.31 or ICD-9/ICD-8:5234 for inclusion criteria [24]. And various genetic principal components were adjusted (age, sex, and 20 others). In this study, data were downloaded from public databases or official websites, all original studies involved have received ethical approval,and the ethical committee approval was not requiredfor this particular aspect of the study.

### Screening of instrumental variable

In this study, SNPs with overlapping druggable genes transcriptional start site (TSS) ± 100KB were extracted from the eQTL dataset as instrumental variables based on the list of druggable genes. Additionally three key assumptions had to be met for MR causal inference. i):IVs must have evidence of characterised exposure, therefore a genome-wide significance threshold (5×10^-8^) and the linkage disequilibrium (LD) threshold R^2^<0.1 were set in this study (based on 1,000 Genomes Project European population) to screen eligible IVs and excluded weak instrumental variables with F-statistics less than 10 [25]; ii):IVs must not be associated with confounders of chronic periodontitis; and iii) IVs must pass the test of horizontal pleiotropy [26-27].

### Statistical analysis

For MR inference methods there are several to pick from; when the number of IVs characterising the exposure is 1, the Wald ratio is chosen as the appropriate method for causal inference [28]; when the number of IVs is greater than or equal to 2, the IVW method with strong statistical validity is chosen. the OR calculation relies on the Wald ratio method [29] and the delta method assesses the corresponding 95% confidence interval. In addition to this, we coordinated the exposure and outcome data according to the effect and non-effect alleles of each SNP. SNPs with allelic mismatches were excluded to ensure data integrity.Finally, FDR was used to adjust for type 1 errors from multiple hypothesis testing (PFDR<0.05) [30].

### Multiple validity testing

Since the IVs used in this MR study originated from single-gene regions, multiple gene region pleiotropy detection methods (e.g., MR-Egger intercept) were no longer applicable, and we used summary-data-based Mendelian randomisation (SMR) and Bayesian co-localisation to complete the single-gene region horizontal pleiotropy association detection, where SMR was combined with Heterogeneity In Dependence Instrumental Variables(HEIDI) to detect heterogeneity in the results [31]. In addition co-localisation of five mutually exclusive models corresponding to five posterior probabilities [32]: PPH0-PPH4. In this study, co-localisation analyses were performed using SNPs within ±100 kb of the TSS of each gene in the eQTL and concluded that the genetic association between the expression level of the gene and the risk of chronic periodontitis was supported by the colocation when PPH4 ≥ 0.6.

### TWAS Validation

FUSION Transcriptome-Wide Association Study (TWAS) analyses are used to predict and test the correlation between gene expression and specific diseases or phenotypes by combining aggregated GWAS statistics with gene expression data, which can be used to predict and test the correlation between gene expression and specific diseases or phenotypes by identifying genes that are significantly correlated with complex traits in individuals who do not have direct measurements of their expression levels, to provide a more comprehensive understanding of the genetic mechanisms of disease and guide future drug development and therapeutic strategies [33-34].

Download the code on Git-hub, containing the relevant code for mapping and making weight files (https://github.com/gusevlab/fusion_twas). top1, blup, lasso, enet, and bslmm are the five prediction models used by FUSION, and we applied FUSION to correlate CP’s genetic effects with eQTL weights to calculate the linear product of Z-values of SNPs and eQTL weights for TWAS analysis.

### Phenome-wide association study

In order to assess the potential side effects of the identified drug targets, we performed PheWAS-MR (https://www.leelabsg.org/resources) on selected phenotype-wide GWAS data (779 in total) from UK Biobank. The original database was adjusted for sample case-control imbalance with cryptic relatedness using the Scalable and Accurate Implementation of Generalised mixed model (SAIGE) (V.0.29) method to account for unbalanced of the case-control ratio [35]. In addition to this, the IVs screening strategy was harmonised with the above and the IVW method was used as the primary method, with MR Egger, Weighted median and Weighted mode as the secondary methods for PheWAS-MR.Finally the FDR (BH) was used to correct for the multiple test results, and a PFDR < 0.05 was considered as the threshold of significance.

## Results

Selected according to the set screening criteria, this study extracted 2288 genes from the eQTL dataset, corresponding to 27,496 IVs, (F-statistics ranging from 29.7-3450.7), and detailed information of the IVs is shown in **Additional Table 1**.

**Table 1.**
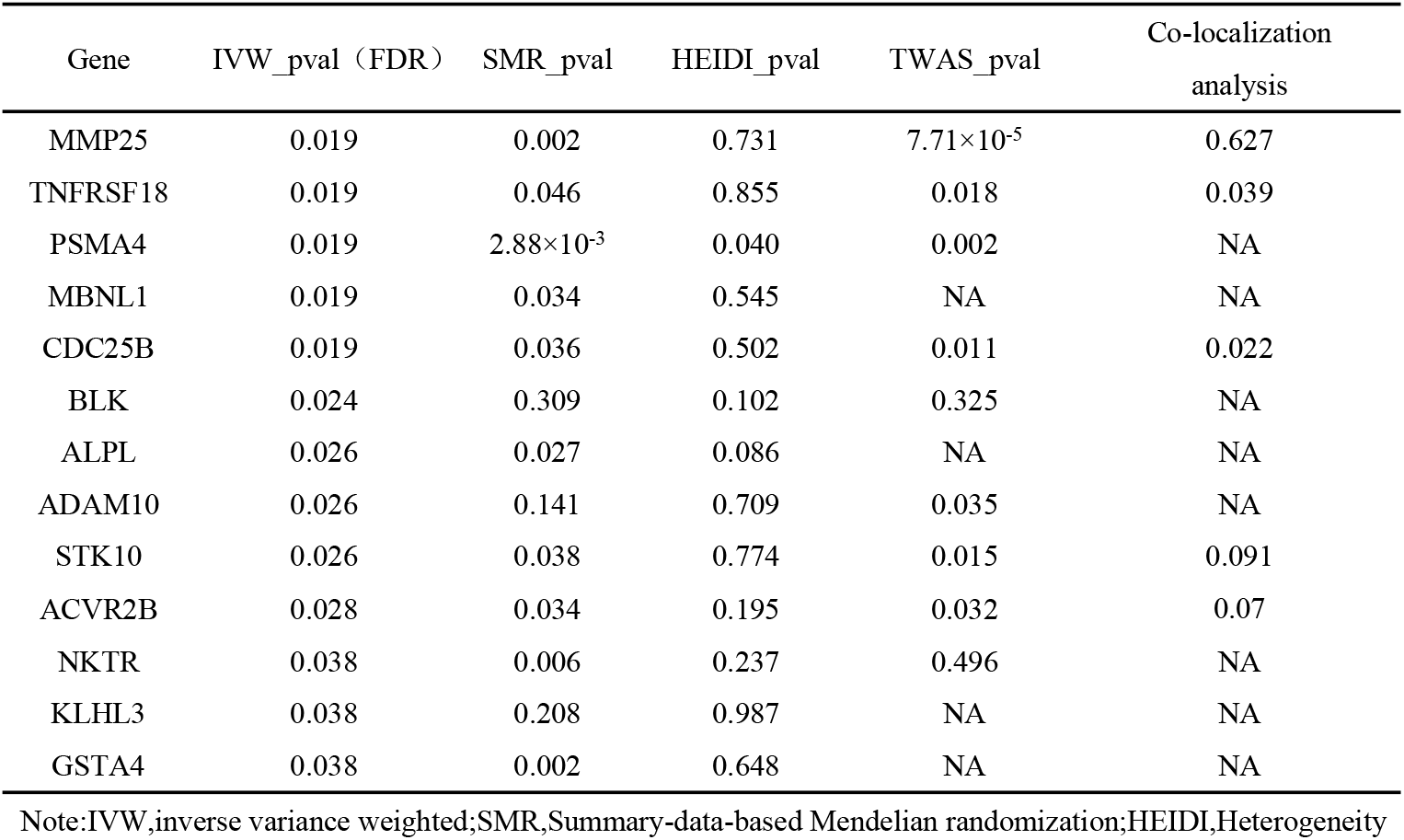
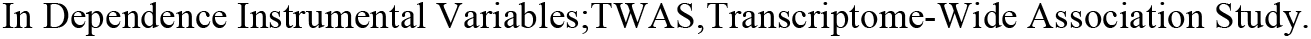
Results of colocalization, SMR and TWAS analysis.

| Gene | IVW_pval (FDR) | SMR_pval | HEIDI_pval | TWAS_pval | Co-localization analysis |
| --- | --- | --- | --- | --- | --- |
| MMP25 | 0.019 | 0.002 | 0.731 | $7.71 \times 10^{-5}$ | 0.627 |
| TNFRSF18 | 0.019 | 0.046 | 0.855 | 0.018 | 0.039 |
| PSMA4 | 0.019 | $2.88 \times 10^{-3}$ | 0.040 | 0.002 | NA |
| MBNL1 | 0.019 | 0.034 | 0.545 | NA | NA |
| CDC25B | 0.019 | 0.036 | 0.502 | 0.011 | 0.022 |
| BLK | 0.024 | 0.309 | 0.102 | 0.325 | NA |
| ALPL | 0.026 | 0.027 | 0.086 | NA | NA |
| ADAM10 | 0.026 | 0.141 | 0.709 | 0.035 | NA |
| STK10 | 0.026 | 0.038 | 0.774 | 0.015 | 0.091 |
| ACVR2B | 0.028 | 0.034 | 0.195 | 0.032 | 0.07 |
| NKTR | 0.038 | 0.006 | 0.237 | 0.496 | NA |
| KLHL3 | 0.038 | 0.208 | 0.987 | NA | NA |
| GSTA4 | 0.038 | 0.002 | 0.648 | NA | NA |

Note: IVW, inverse variance weighted; SMR, Summary-data-based Mendelian randomization; HEIDI, Heterogeneity
In Dependence Instrumental Variables;TWAS,Transcriptome-Wide Association Study.

This study ultimately investigated the genetic association of 2288 potential drug genes with CP using MR (**Additional Table 2**), and after FDR correction, 13 genes were found to be genetically associated with CP (**Fig. 2A and Fig. 2B**), including matrix metallopeptidase 25 (MMP25) [odds ratio (OR)=1.13, 95% CI: 1.07-1.19, P = 1.45 × 10^-5^], proteasome 20S subunit alpha 4 (PSMA4) (OR = 1.39, 95% CI: 1.19-1.62, P = 2.74 × 10^-5^) and muscleblind like splicing regulator 1 (MBNL1) (OR=1.17, 95% CI: 1.08-1.26, P =3.70×10^-5^) and seven other drug gene expression levels were positively correlated with CP, whereas the expression levels of TNF receptor superfamily member 18 (TNFRSF18) (OR=0.81, 95% CI. 0.73-0.89, P = 2.01 × 10^-5^), serine/threonine kinase 10 (STK10) (OR = 0.78, 95% CI: 0.69-0.89, P = 1.02 × 10^-4^) and activin A receptor type 2B (ACVR2B) (OR = 0.85, 95% CI: 0.79-0.93, P = 1.21 × 10^-4^) and eight other drug gene expression levels were negatively correlated with CP.

**Figure 2.**
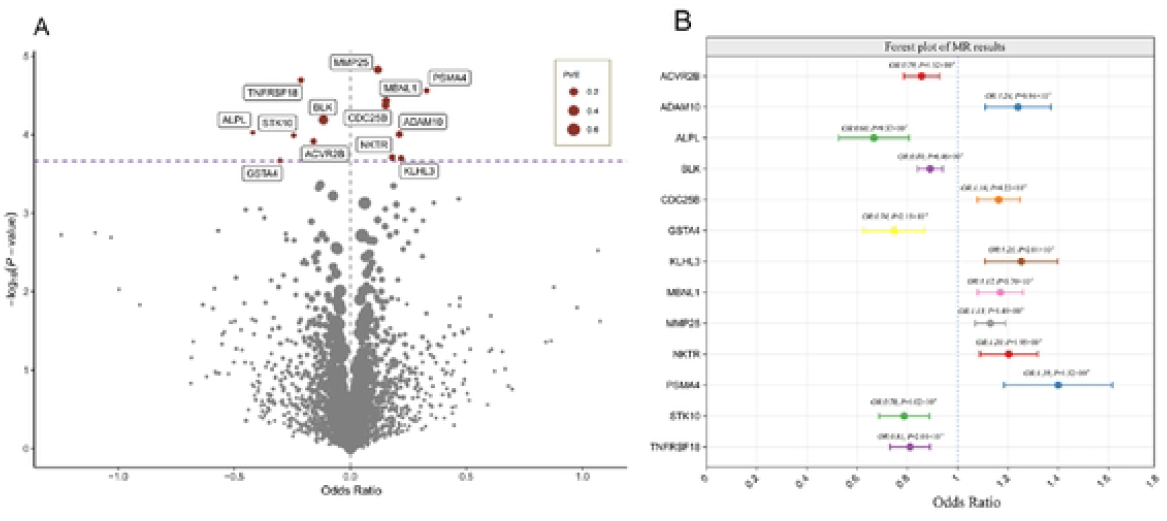
Mendelian randomisation results of druggable genes with CP. A:Volcano plot of MR results. The size of the dots represents the size of PVE, and the purple dotted line indicates the threshold after FDR<0.05 correction.PVE: proportion of variance explained; PVE=sum((beta.exposure^2)/(beta.exposure^2 + samplesize.exposure *se.exposure^2). B: Forest plot of MR results.

Multiplicity testing showed that only MMP25 was supported by SMR&HEIDI results and co-localisation results, and passed the validated Fusion TWAS test. In addition, TNFRSF18, cell division cycle 25B (CDC25B), STK10, and ACVR2B passed the SMR&HEIDI and validated Fusion TWAS tests, but were not supported by co-localisation results (see **Table 1** for detailed results). Unfortunately, PSMA4 and ADAM10 only passed the validated Fusion TWAS test.

Then, based on the MR, SMR&HEIDI, co-localisation and validated Fusion TWAS test results, we assigned endorsement grades to CP-associated pharmacogenes, among which we maintained the highest confidence in MMP25, as the MR, SMR&HEIDI, co-localisation, and validated Fusion TWAS test results all provided sufficient evidence of genetic linkage with CP genetic association. We also maintained some confidence in TNFRSF18, CDC25B, STK10, and ACVR2B as they passed MR, SMR&HEIDI, and validated Fusion TWAS test results, and just failed co-localisation, which may be due to the low number of risk loci in the raw GWAS data for CP. We had lower confidence in eight drug genes such as PSMA4, ADAM10 and NKTR.

Finally, since drugs usually act via blood circulation, we used PheWAS-MR to evaluate the potential side effects of MMP25 as a drug target (**Fig. 3**). IVW results showed (FDR<0.05), MMP25 was associated with 19 phenotypes (**Additional Table 3**), where MMP25 was the Premature beats (OR=1.472 (OR=1.472, P=3.77×10^-6^), Dizziness and giddiness (OR=1.74, P=6.97×10^-6^) and Type 2 diabetes with neurological manifestations (OR=1.39, P=9.91×10^-6^) were 13 phenotypic risk Factors. In contrast, MMP25 was a risk factor for Otitis externa (OR=0.717, P=2.42×10-5), Other disorders of metabolism (OR=0.834, P=1.41×10^-4^) and Cellulitis and abscess of trunk (OR=0.733, P =2.68×10^-4^) and other protective factors for six phenotypes.

**Figure 3.**
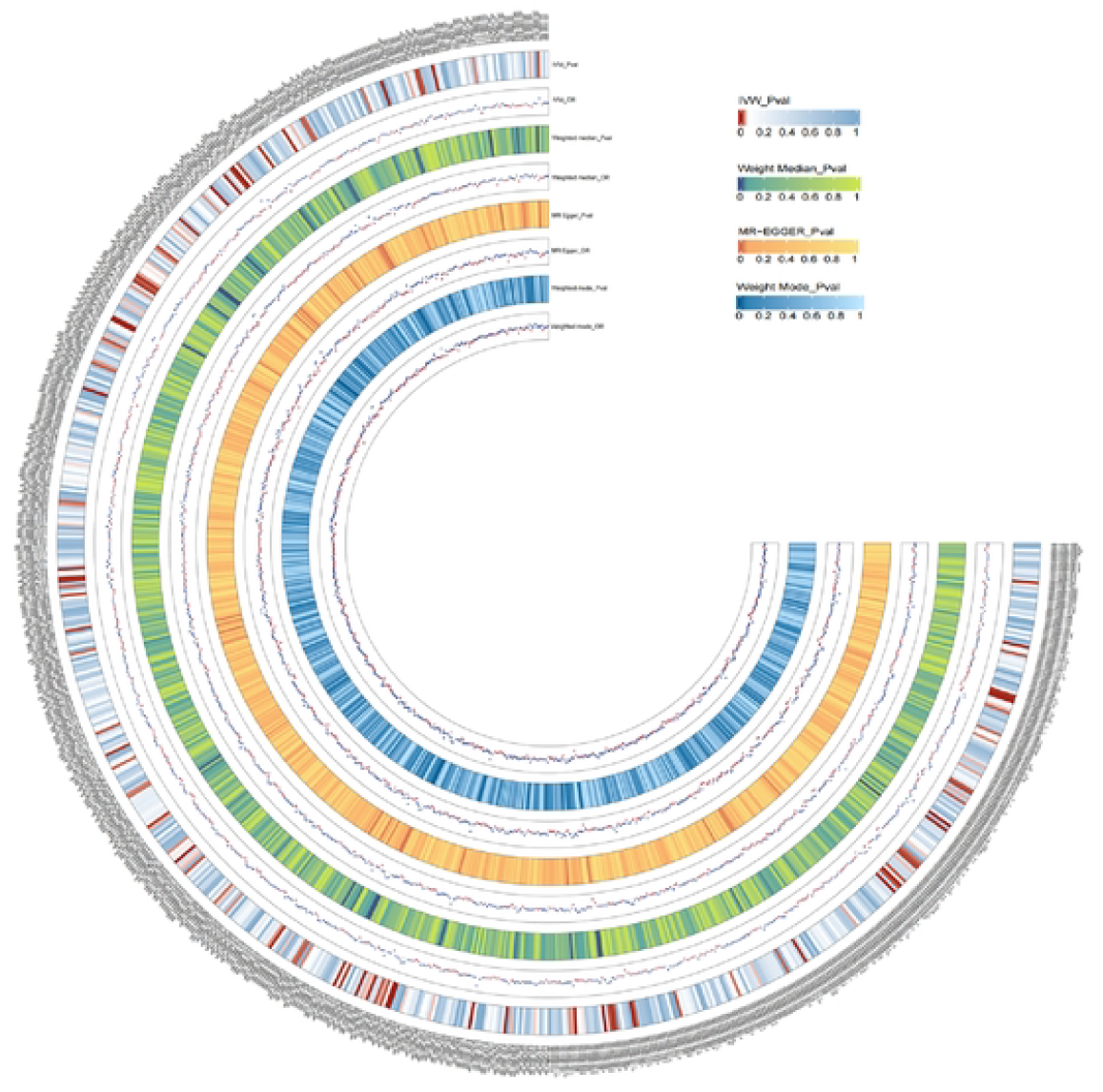
Results of MR studies of the full phenotypic range of MMP25.

The main statistical analysis was performed in R (version 4.3.3). Using the “coloc” software package (version 5.2.1) for Bayesian colocation analysis, Two-sample MR analysis was conducted with “MendelianRandomization” (version 0.6.6). SMR analysis from https://yanglab.westlake.edu.cn/software/smr/#Download. TWAS analysis code from https://github.com/DelinLi/TWAS.

## Discussion

This study used TSMR, Fusion TWAS, SMR, and co-localisation a range of methods to assess the genetic association between druggable targets and CP. In conclusion, MMP25 passed all the tests and therefore we maintain maximum confidence in it, in addition to TNFRSF18, CDC25B, STK10 and ACVR2B. Finally, we investigated the potential side effects of MMP25 as a target using PheWAS-MR.

MMP25 is an enzyme belonging to the MMP family.MMP participates in many physiological processes and is a group of highly conserved endopeptidases [36-37]. Numerous researches have indicated that the MMP family was vital in driving inflammation in periodontitis, promoting periodontal tissue destruction through degradation and remodelling of extracellular matrix components. Some in vitro experiments verified that the pathogenic bacterium causing CP (Fusarium nucleatum) promotes further inflammation in periodontal tissues, including alveolar bone resorption and cellular matrix degradation, by stimulating macrophages to increase the secretion of MMP9 [38]. It has been demonstrated that there may be a positive feedback loop between MMP13 and MMP9, which together promote periodontal tissue inflammation and bone loss [39]. Initially, it was found that MMP25 was specifically expressed in peripheral blood leukocytes and upregulated in mouse models of cancer and infection, and the close relationship with periodontitis has been repeatedly demonstrated, for example, a randomised controlled trial found that MMP25 levels in gingival sulcus fluid of patients with chronic periodontitis were higher than in healthy controls [40],Another basic study found that levels of MMP25 in periodontal tissues of chronic periodontitis increased. In conclusion, MMP25 is closely related to chronic periodontitis, which is consistent with the conclusions reached in the current study [41].

TNFRSF18 is also referred to as Glucocorticoid-Induced TNFR-Related protein (GITR), which has been linked to immune system homeostasis and inflammatory disease development. Currently, although there are no direct studies on TNFRSF18 and CP, it has been reported that a TNFRSF member, TNFSF14, was important in promoting the CXCL10 secretion and CXCL11 from human gingival fibroblasts, the infiltration of Th1 cells into foci of inflammatory illness can be promoted, and was a prospective target for periodontitis treatment.

STK10 gene belongs to Ste20 family and encodes a serine/threonine protein kinase. The protein binds to and phosphorylates polo-like kinase 1, its kinase-dead overexpression disturbs cell cycle progression [42-43]; CDC25B belongs to phosphatases CDC25 family, and is necessary for cells to enter mitosis [44]; ACVR2B acts as a signalling mediator with the ability to transmit activin signals that regulate many pathological and physiological processes [45-46]. STK10, CDC25B and ACVR2B have been little studied in CP, so further experiments are needed to verify their association with CP.

In this study, we used many effective methods to improve the robustness of the MR study; firstly: the included instrumental variables all have F-statistics greater than 10, so they have the ability to characterise the drug genes; secondly, in order to unify the genomic co-ordinates, we localised the current MR study using SNPs to the human genome HG19/GRCh37; thirdly, we used Fusion TWAS to enhance the association between the drug association between genes and CP; fourth, all GWAS data were from European ethnicity to reduce genetic confounding due to ethnic differences; fifth, we selected only SNPs within the cis-eqtl region and used SMR and co-localisation to avoid horizontal pleiotropy.

There are still limitations in this study. First, although MR verified the causal relationship between drug gene targets and CP genetically via the IV method, the MR approach assumes a linear relationship between CP and drug gene expression levels. Therefore, MR results should be viewed with caution in case of nonlinear correlation between outcome and exposure. Secondly, the GWAS data of the current study on the association between drug genes and CP were obtained from people of European descent. Therefore, the MR results need to be further investigated and validated when extended to other ethnic groups to ensure wider applicability. Thirdly, this study only concentrated on the causality between the pharmacogenetic cis-eQTL and CP, and lacked the study of trans-eQTL. Finally, the results of this study need to be supported by clinical data, which is also the direction of our future efforts.

## Conclusion

Five potential CP drug targets were identified in this study using TSMR, Fusion TWAS, SMR, and a series of co-localisation methods, of which MMP25 passed all the tests, so we maintain the utmost confidence in it. The findings provide a rationale for future CP drug development and reduce the cost of drug development to some extent.

## Data Availability

All relevant data are within the manuscript and its Supporting Information files.

https://www.leelabsg.org/resources.

https://github.com/gusevlab/fusion_twas

https://www.finngen.fi/fi

https://eqtlgen.org/

## Supporting information

**S1 Fig.Flowchart of the design of this study**.

**S2 Fig.Mendelian randomisation results of druggable genes with CP**.

**S3 Fig.Results of MR studies of the full phenotypic range of MMP25**.

**S1 Table. Details of extracted instrumental variables**.

**S2 Table.MR results**.

**S3 Table.PheWAS-MR results**.

**S1 Checklist.STROBE-MR-checklist**.

## Acknowledgments

This study would like to thank the official eQTLGen (https://eqtlgen.org/) website and the Finnish database version R11 (https://www.finngen.fi/fi) and public catalog GWAS data provided.Thanks to all authors for their contributions.

## Author Contributions

**Data curation:**Zhong-wei Cheng,Fang Wang,Shen-hu Liang.

**Formal analysis:**Zhong-wei Cheng,Fang Wang,Qing-gao Song.

**Project administration:**Qing-gao Song.

**Software:**Mei-mei Ran.

**Supervision:**Shen-hu Liang.

**Writing–original draft:**Zhong-wei Cheng.

## Data availability statement

All data included in this study are available upon request through contact with the corresponding author.

## Ethics statement

In this study, data were downloaded from public databases or official websites, all original studies involved have received ethical approval.

